# Assessing knowledge, attitudes, and behaviours toward salt and sugar consumption in the Central Division of Fiji

**DOI:** 10.1101/2024.06.14.24308937

**Authors:** Gade Waqa, Colin Bell, Joseph Alvin Santos, Kris Rogers, Anasaini Moala Silatolu, Erica Reeve, Aliyah Palu, Alvina Deo, Jacqui Webster, Briar McKenzie

**Affiliations:** Pacific Research Centre for the Prevention of Obesity and Non-communicable Diseases (C-POND), Fiji Institute of Pacific Health Research, Fiji National University, Suva, Fiji; Institute for Health Transformation, Global Centre for Preventive Health and Nutrition, Deakin University, Geelong, Australia; The George Institute for Global Health, University of New South Wales, Australia; Department of Business Economics, Health and Social Care (DEASS); University of Applied Sciences and Arts of Southern Switzerland; Manno, Ticino, Switzerland; Faculty of Health, The University of Technology Sydney, Australia; Ministry of Health, Suva, Fiji

## Abstract

**Background:** Excessive salt and sugar intake influence the global burden of non-communicable diseases. This study aimed to describe the knowledge, attitudes and behaviours (KAB) of Fijian adults relating to salt and sugar consumption to inform policy interventions in Fiji.

**Methods:** A randomised stratified sample of 700 adults in the Central Division of Fiji were selected. Questions on salt-related KAB were adapted from the World Health Organization Noncommunicable Disease Risk Factor survey, and questions on sugar were developed following a similar structure. Locally trained research assistants collected data. For analyses, population and sample weights were applied, and difference between predefined subgroups (sex, age, ethnicity and area of residence) were compared using weighted chi-square tests.

**Results:** 534 adults participated (response rate,76%). Over 80% of participants (82% (95% CI 78.5 to 84.9%)) identified that consuming too much salt or salty sauce can lead to hypertension. More than 90% recognized that consuming too much sugar can lead to diabetes (92.3% (89.7 to 94.3%)). Approximately 80% of participants thought it was somewhat or very important to lower salt and sugar intake in their diet (79.8% (95%CI, 76.1 to 83.0) and 84.2% (80.8 to 87.1%), respectively). However, self-reported behaviours did not align, with almost 40% adding salt or salty sauces as standard practice when cooking (37.3% (32.7 to 42.2%)) and 65% (60.6 to 68.5%) reporting that they add sugar to drinks daily. Younger compared to older individuals (18 to 44 years vs 45 years and older) and men compared to women, had lower levels of KAB.

**Conclusion:** Despite having knowledge of the health impacts of consuming excess salt and sugar and positive attitudes towards reducing consumption, many people reported behaviours likely to contribute to high salt and sugar intake. These findings highlight the need for interventions that incentivise healthier choices, through behaviour change communications and the creation of supportive food environments.

## Introduction

Non-communicable diseases (NCDs) account for approximately 74% of all deaths globally,^1^ with cardiovascular disease accounting for most NCD deaths. The global burden of NCDs is influenced by dietary factors, particularly excessive salt and sugar intake^2 3^. High salt consumption is linked to elevated blood pressure and increased risks of CVDs such as heart disease and stroke^4 5^. Excessive sugar intake, especially from sugar-sweetened beverages (SSBs), is associated with various health issues, including obesity and diabetes^6-10^.

The shift from traditional diets based on locally sourced foods to diets dominated by imported and processed foods has exacerbated the problem of diet-related diseases in the Pacific Islands^11-13^. Evidence suggests an association between the dietary transition to a higher population intake of salt and sugar with the rising prevalence of NCDs^13 14^ In 2017, WHO identified reducing sugar and salt among the best ways to prevent and control NCDs^3^. In response, Pacific countries proposed maximum acceptable regional targets for salt in eight selected food categories ^15^. However, this initiative was voluntary, and coordinated efforts to reduce salt intake at the population level have been limited^15 16^ leading to ineffective interventions^17^. While the Pacific region, including Fiji, has implemented fiscal policy interventions for SSBs^18 19^ in line with WHO recommendations on “best buys^3^ these policies have not been fully aligned with WHO guidelines and require strengthening to achieve health benefits^18^.

We have previously identified that salt intake was almost double the maximum amount and sugar intake three times the ideal amount, as recommended by the WHO, highlighting the need for interventions to reduce intake in Fiji^20^. To better understand what interventions are needed, it is essential to assess people’s knowledge, attitudes, and behaviours (KAB) toward salt and sugar consumption. Therefore, this study aimed to describe the knowledge, attitudes and behaviours, relating to salt and sugar consumption, of a representative sample of adults in the Central Division of Fiji.

## Materials and methods

This cross-sectional survey was part of a broader study to strengthen food policies in Fiji^21^. The objective of the larger project was to identify factors needed to achieve effective food policy implementation for healthier food environments in the Pacific. The survey aimed to assess salt and sugar intake, salt and sugar-related KABs, food security, and the impact of COVID-19. Findings on salt and sugar intake, food security, and the effects of COVID-19 have been published, with methods for these survey components explained in respective papers^20 22^. Ethics approval for this survey was obtained from the University of New South Wales (HC200469) and the Fiji National University College of Human Health Research Ethical Committee (CHHREC264.20).

### Sample size and recruitment

Two enumeration areas within the Central Division of Fiji were randomly selected for the survey: Waidamudamu Medical Zone (an urban area) and Deuba Medical Zone (a rural area). A randomized stratified sampling approach was employed, considering age, sex, and ethnicity to ensure representation from demographic groups. The sample size was calculated based on the primary objective of the survey, which was to act as a baseline measure for monitoring changes in salt and sugar intake. Based on an estimated 16% non-response rate, 700 individuals (350 for each area) were sampled^20^. This sample size was determined to achieve at least 80% power to detect specific changes in salt intake (0.6g/day with a standard deviation of 3.6) and sugar intake (0.9 absolute percent change with a standard deviation of 5.4) following the intervention with a precision (95% confidence interval) of 7.8%.

Permission to enter villages and conduct the study was gained through the Ministry of iTaukei Affairs of the Fiji government, with relevant approvals obtained from the Provincial Council office and the Nasinu and Nausori Municipalities under the Ministry of Local Government. Traditional Fijian customs, such as the “i sevusevu” kava ceremony, were performed to seek approval from local village chiefs, leaders of faith-based organizations and selected community members. Information sessions were conducted at neutral locations such as village meeting halls, within each village, to inform residents about the survey and address any questions or concerns. Research assistants visited the homes of selected participants, invited them to participate in the survey, and provided them with information about the study. If no one aged 18 or above was at home, then a repeat visit was made at a later stage. Participant information sheets and consent forms were available in English, Hindi, and Fijian, and oral translations were provided as needed. Eligibility was based on age (aged 18 years or older), and the ability to provide written informed consent. Surveys were predominantly conducted on weekdays (Monday – Friday), with arrangements made for weekend data collection as necessary to accommodate participants’ schedules. Repeat visits were made to households if initial attempts to contact residents were unsuccessful or only individuals under 18 were present. The surveys took place from the first week of March to the last week of June 2022.

### Survey instrument, salt and sugar-related knowledge, attitudes and behaviours

The salt and sugar questions used were based on the WHO STEPS survey for salt, with a similar structure followed to develop a set of sugar-specific questions ^23^. Prior to implementation, the research team piloted the tool to ensure its comprehensiveness and clarity within the research team. See **supplementary material** for the KAB questionnaire.

Dietary salt, according to the WHO STEPS survey questionnaire, was defined as ordinary table salt, unrefined salts such as sea salt, iodized salt, and salty sauces such as soy sauce or ketchup. Dietary sugar was defined as raw and white sugar (including sugar lumps), brown sugar, cane sugar, caster, and icing sugar.

#### Knowledge

Participants were assessed on their knowledge of salt and sugar through three questions each, including knowledge of the recommended daily intake amount, recognition of teaspoon equivalents for the recommended intake, and understanding of the health implications of excessive salt or sugar consumption, with response options indicating various health conditions (e.g. hypertension, diabetes) or “don’t know”.

#### Attitudes

Attitudes towards salt and sugar intake were evaluated using single questions each, asking participants to rate the importance of reducing intake in their diet on a 5-point Likert scale ranging from “very important” to “not at all important” or “don’t know.”

#### Behaviours

Salt and sugar-related behaviours were assessed through six questions each. For salt-related behaviours, inquiries included discretionary salt use during cooking and meal consumption; frequency of consuming processed foods high in salt (such as packaged salty snacks, canned salty food including pickles and preserves, salty food prepared at a fast-food restaurant, cheese, and processed meat); efforts to reduce salt intake, and specific actions taken to achieve this reduction (e.g., limit consumption of packaged processed foods, limit consumption of takeaways/fast food, look at the salt/sodium content on food labels, buy low salt alternatives, use spices other than salt when cooking or other).

For sugar-related behaviours, questions addressed the frequency of consuming drinks with added sugar (hot or cold e.g. coffee, tea, milo/hot chocolate, water, juice); the amount of sugar added to drinks; consumption of sugar-sweetened beverages (e.g. fizzy drinks, sodas, juice, raro/concentrate); efforts to reduce sugar intake, and specific actions taken to achieve this reduction (e.g., limit consumption of packaged processed foods, limit consumption of sugar-sweetened beverages, limit the addition of sugar to hot or cold drinks, limit the use of instant drink mixes (e.g., coffee mixers), limit consumption of confectionary, limit consumption of baked goods, like cakes and sweet biscuits, sweet pastries, and ice cream, buy low sugar alternatives, other).

Responses for all behaviour -related questions ranged from specific frequency options (e.g, “always” to “never”) to categorical choices regarding efforts to reduce intake and specific behavioural actions undertaken.

### Data collection

The data collection process involved conducting interviews with participants in their homes or other convenient spaces, each lasting approximately an hour. Locally trained research assistants collected data using a mobile Android Package Kit (APK) containing the 19 multiple-choice questions designed to assess salt and sugar-related KAB. In addition to the KAB, demographics, health status, and physical measurements were collected. Participant demographic data included sex, age, ethnicity, highest education level, and household living arrangement (living alone vs sharing/living with others). Physical measurements, including height, weight, waist circumference, and three blood pressure readings, were taken. Further information on collecting demographics and physical measurements have been published previously ^20 22^.

### Data analysis

All data were analyzed using STATA BE V17.0 for Windows (Stata Corp LP, College Station, TX, USA). Analyses were weighted to reflect the probability based on the random selection of individuals (sample weight) and to match the appropriate estimates of the population in Deuba and Waidamudamu (population weight). The differences between subgroups (by sex (female, male), age (18-44, 45 years and older), ethnicity (iTaukei or Indigenous Fijians, Fijian of Indian descent or other), and area (Waidamudamu Medical Zone, Medical Zone) were compared using weighted chi-square tests. For all analyses, the svy command in Stata accounted for strata effects, and the Taylor linearization method was employed for variance estimation. Results were reported as mean (for continuous variables) or proportion (for categorical variables) with standard error (SE) or 95% confidence interval (CI) as appropriate.

## Results

### Demographics and health status

A total of 534 people completed the survey (response rate 76%). Half of the population were female (50.4%), and the mean age of the study population was 42 years. Most participants had secondary education (69.4%) and lived with others (95.8%). Just under half of the population stated they were iTaukei (Indigenous) Fijians (46.3%).

Most respondents (approximately 90%) reported that their health was either good, very good, or excellent. However, a third reported being current smokers (28.7% (95% CI, 25.4 to 32.3)), 28% reported they had been diagnosed with high blood pressure (28.0% (24.5 to 31.7)) and 10% reported having a history of diabetes (9.7% (7.7 to 12.2)). Additionally, more than half of the participants were classified as hypertensive based on blood pressure measures taken during the survey 50.8% (46.8 to 54.8%). Mean body mass index was 28.8 kg/m^2^ (28.2 to 29.3), with a higher prevalence of obesity observed among women (50.2% (44.6 to 55.8)) compared to men (32.5% ((27.2 to 38.3)). Characteristics of nutrition survey participants, anthropometric measurements and self-reported health status have been previously published^20^.

### Salt

#### Salt-related knowledge

Most participants were aware of the relationship between high salt intakes and the increased risk of hypertension (82% (95% CI, 78.5 to 84.9)), however just over a quarter (27% (95%CI,23.8 to 31.4)) were aware of the relationship between high salt intake and the risk of having a stroke. Only 16% of participants knew that the maximum recommended amount of salt per day is 5g or less (16.2% (13.4 to 19.5%)), of which 25% (25.1% (21.7 to 28.8%)) knew that this looked like one teaspoon of salt. Knowledge differed by age, area, and ethnicity. For example, in regard to knowledge of the recommended daily intake of salt, significantly more Fijians of Indian descent and other ethnicities compared to iTaukei Fijians were aware of the recommendations (21.3% (16.7% vs 25.9%) vs. 10.3% (6.4% vs 14.2%)). There was a similar difference in awareness between those in a rural setting (Deuba) compared to urban (Waidamudamu) (21.4% (16.6 to 26.1%) vs 12.8% (8.8 to 16.7%)), **table 1**.

**Table 1:**
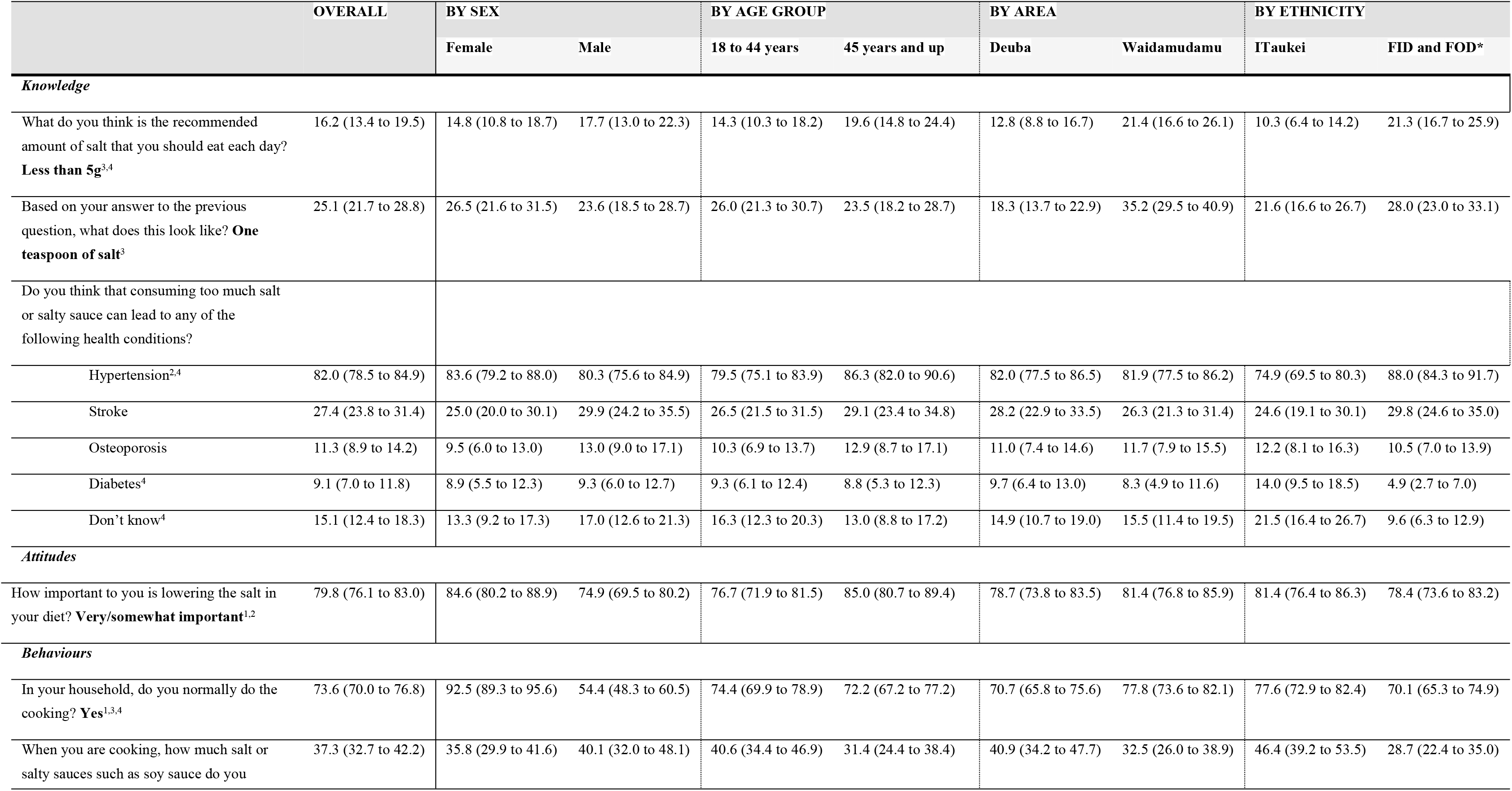
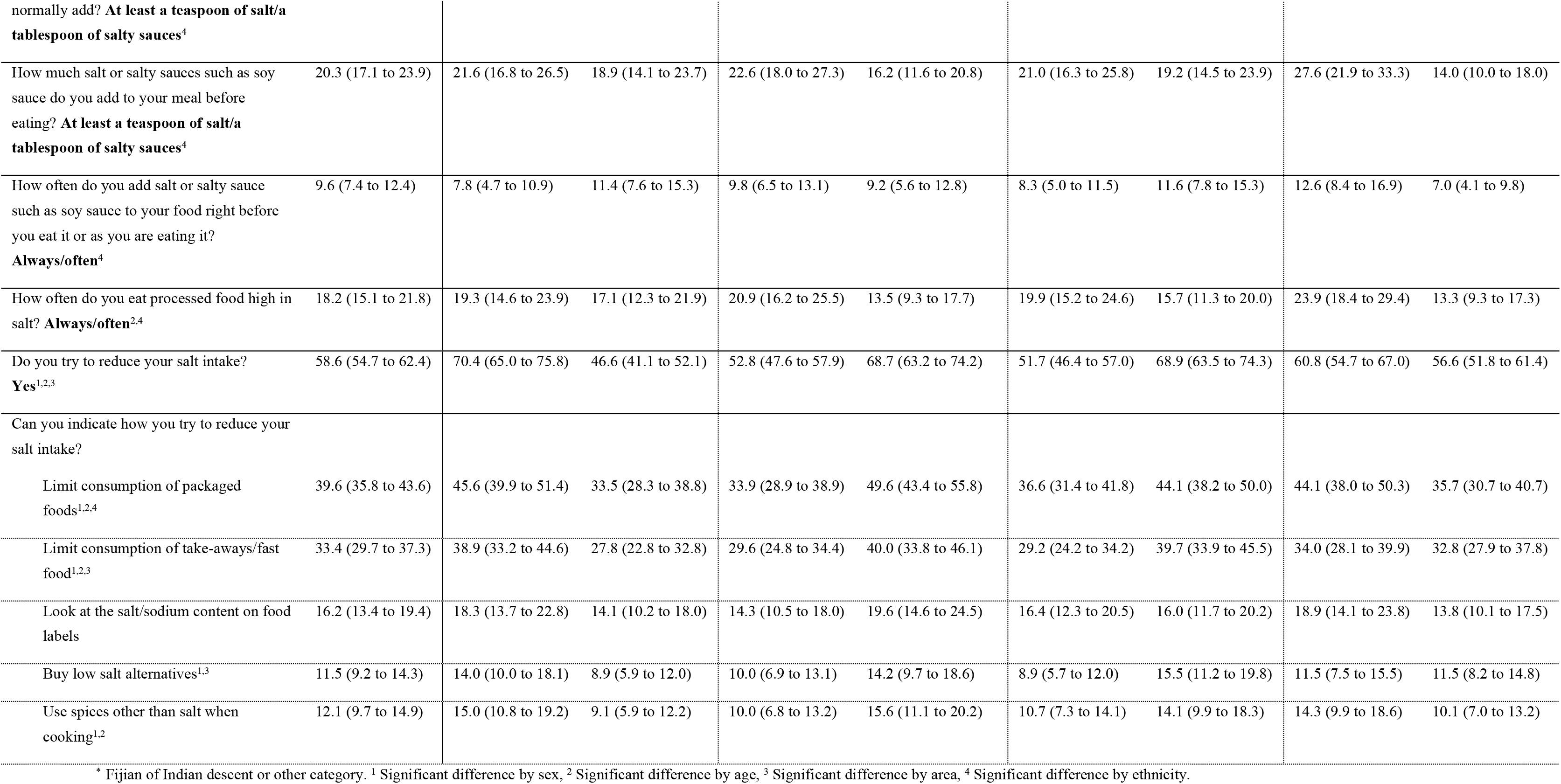
Knowledge, attitudes, and behaviours towards salt overall and by subgroups (weighted %, 95% CI)

### Salt-related attitudes

Seventy-nine percent of participants (79.8% (95%CI, 76.1 to 83.0)) said it was very or somewhat important to lower the salt in their diet differing by sex and age, with higher importance reported by females than males (84.6% (80.2 to 88.9) vs 74.9% (69.5 to 80.2%)) and for those in the older age range vs younger (85% (80.7 to 89.4) vs 76.7% (71.9 to 81.5%)),

### Salt-related behaviours

Almost three-quarters (73.6% (70.0 to 76.8%)) of respondents said they were the main cook at home, with 93% of surveyed women reporting that they were the main cook (92.5% (89.3 to 95.6%)). Just over a third reported that they add at least a teaspoon of salt or a tablespoon of salty sauces normally when they are cooking (37.3% (32.7 to 42.2%)), and almost 10% reported adding salt or salty sauce either always or often right before eating (9.6% (7.4 to 12.4%)). More than half of the study population (58.6% (54.7 to 62.4%)) reported trying to reduce their salt intake, with the most common behaviours to reduce salt intake being limiting the consumption of packaged foods (reported by 39.6% (35.8 to 43.6%)), **table 1**.

There were key differences in salt-related behaviours by subgroups (**table 1**), for example, a higher proportion of iTaukei Fijians reported adding a teaspoon of salt or a tablespoon of salty sauces when cooking (46.4 (39.2 to 53.5%) vs. 28.7% (22.4 to 35.0%)) and always or often adding salt or soy sauce when they eat (12.6% (8.4 to 16.9%) vs. 7.0% (4.1 to 9.8%)), compared to Fijians with Indian descent and other. Further, significantly more women compared to men reported trying to reduce their salt intake (70.4% (65.0 to 75.8%) vs 46.6% (41.1 to 52.1%)), similarly a higher proportion of older people reported trying to reduce their salt intake (68.7% (63.2 to 74.2%) vs 52.8% (47.6 to 57.9%) in the younger age group), and more people from the urban area compared to rural (68.9% (63.5 to 74.3) vs. 51.7% (46.4 to 57.0%)).

#### Sugar

##### Sugar-related knowledge

Most participants were aware of the relationship between high intakes of sugar and diabetes (92.3% (95% CI 89.7 to 94.3%); however, the relationship between high sugar intake and the risk of poor dental health (36% (26.9 to 34.6%)), and obesity (27.9% (24.2 to 31.8%)) were less well known. Only 22% (18.6 to 25.7%) of participants knew the recommended daily sugar intake of less than 10% of total energy intake (**Table 2**). The level of sugar-related knowledge was similar across subgroups, except for awareness around the relationship between sugar intake and poor dental health, which differed by area (35.8% (30.2 to 41.3%) urban vs 27.2% (22.0 to 32.4%) rural) and ethnicity (35.7% (30.3 to 41.1%) Fijian of Indian or other descent, vs 24.7% (19.3 to 30.1%) for iTaukei Fijians).

**Table 2.** Knowledge, attitudes, and behaviours towards sugar overall and by subgroups (weighted %, 95% CI)

|  | OVERALL | BY SEX |  | BY AGE GROUP |  | BY AREA |  | BY ETHNICITY |  |
| --- | --- | --- | --- | --- | --- | --- | --- | --- | --- |
|  |  | Female | Male | 18 to 44 years | 45 years and up | Deuba | Waidamudamu | ITaukei | FID and FOD* |
| Knowledge |  |  |  |  |  |  |  |  |  |
| What do you think is the recommended amount of sugar that you should eat each day? **Less than 10% of total energy intake** | 22.0 (18.6 to 25.7) | 19.5 (14.9 to 24.1) | 24.4 (19.1 to 29.8) | 22.2 (17.5 to 26.9) | 21.5 (16.3 to 26.7) | 19.6 (14.8 to 24.3) | 25.5 (20.3 to 30.7) | 19.8 (14.7 to 24.9) | 23.8 (18.9 to 28.7) |
| Do you think that consuming too much sugar or sugar sweetened beverage can lead to any of the following health conditions? |  |  |  |  |  |  |  |  |  |
| Diabetes | 92.3 (89.7 to 94.3) | 92.0 (88.7 to 95.2) | 92.6 (89.4 to 95.8) | 92.1 (89.1 to 95.1) | 92.6 (89.2 to 96.0) | 91.9 (88.7 to 95.1) | 92.9 (89.8 to 96.0) | 90.6 (87.0 to 94.2) | 93.7 (90.8 to 96.6) |
| Obesity | 27.9 (24.2 to 31.8) | 30.6 (25.2 to 36.1) | 25.1 (19.8 to 30.3) | 29.6 (24.6 to 34.7) | 24.8 (19.3 to 30.3) | 28.1 (22.8 to 33.4) | 27.5 (22.2 to 32.7) | 27.9 (22.1 to 33.6) | 27.8 (22.8 to 32.9) |
| Heart disease | 20.8 (17.6 to 24.5) | 20.8 (16.1 to 25.6) | 20.8 (15.9 to 25.8) | 19.5 (15.1 to 24.0) | 23.1 (17.7 to 28.5) | 21.9 (17.1 to 26.6) | 19.3 (14.6 to 24.1) | 22.4 (17.2 to 27.7) | 19.5 (15.0 to 24.0) |
| High cholesterol | 15.9 (13.1 to 19.2) | 16.3 (12.0 to 20.6) | 15.4 (11.1 to 19.8) | 14.8 (10.9 to 18.7) | 17.8 (12.8 to 22.7) | 14.4 (10.4 to 18.5) | 18.1 (13.5 to 22.7) | 14.7 (10.2 to 19.1) | 16.9 (12.7 to 21.1) |
| Poor dental health<sup>3,4</sup> | 30.6 (26.9 to 34.6) | 29.4 (24.2 to 34.5) | 31.9 (26.2 to 37.6) | 28.6 (23.6 to 33.6) | 34.1 (28.3 to 40.0) | 27.2 (22.0 to 32.4) | 35.8 (30.2 to 41.3) | 24.7 (19.3 to 30.1) | 35.7 (30.3 to 41.1) |
| Don't know | 4.8 (3.2 to 7.0) | 5.4 (2.6 to 8.2) | 4.1 (1.7 to 6.6) | 5.1 (2.7 to 7.5) | 4.2 (1.4 to 7.0) | 4.6 (2.1 to 7.1) | 5.0 (2.4 to 7.7) | 4.8 (2.1 to 7.6) | 4.8 (2.3 to 7.3) |
| Attitudes |  |  |  |  |  |  |  |  |  |
| How important to you is lowering the sugar in your diet? **Very/somewhat important**<sup>1,2</sup> | 84.2 (80.8 to 87.1) | 89.4 (85.8 to 93.0) | 78.9 (73.8 to 84.0) | 81.6 (77.2 to 85.9) | 88.8 (84.9 to 92.8) | 82.4 (77.9 to 86.9) | 87.0 (83.0 to 90.9) | 85.1 (80.6 to 89.6) | 83.5 (79.2 to 87.8) |
| Behaviours |  |  |  |  |  |  |  |  |  |
| How often do you drink drinks that you add sugar to? **Daily/5-6 days per week**<sup>1,4</sup> | 64.6 (60.6 to 68.5) | 60.1 (54.4 to 65.8) | 69.3 (63.9 to 74.7) | 66.1 (61.0 to 71.3) | 62.1 (56.0 to 68.1) | 66.0 (60.6 to 71.4) | 62.6 (57.0 to 68.2) | 71.0 (65.5 to 76.5) | 59.2 (53.5 to 64.8) |
| How often do you drink sugar sweetened beverages? **Daily/5-6 days per week**<sup>1,2</sup> | 19.9 (16.8 to 23.5) | 16.5 (12.2 to 20.7) | 23.5 (18.3 to 28.7) | 24.8 (19.9 to 29.6) | 11.5 (7.9 to 15.0) | 17.8 (13.2 to 22.3) | 23.2 (18.4 to 28.0) | 19.2 (14.2 to 24.2) | 20.6 (16.1 to 25.1) |
|  |  | Female | Male | 18 to 44 years | 45 years and up | Deuba | Waidamudamu | ITaukei | FID and FOD* |
| Do you try to reduce your sugar intake?<br>Yes<sup>1,2,3</sup> | 65.1 (61.0 to 69.0) | 73.5 (68.2 to 78.8) | 56.6 (50.6 to 62.5) | 59.7 (54.3 to 65.1) | 74.5 (69.0 to 80.0) | 60.2 (54.6 to 65.8) | 72.4 (67.2 to 77.7) | 64.8 (58.8 to 70.8) | 65.3 (60.1 to 70.6) |
| Can you please indicate how you try to<br>reduce your sugar intake? |  |  |  |  |  |  |  |  |  |
| Limit consumption of packaged<br>processed foods<sup>2</sup> | 31.9 (28.1 to 35.9) | 33.9 (28.5 to 39.3) | 29.8 (24.3 to 35.4) | 27.6 (22.6 to 32.6) | 39.5 (33.3 to 45.6) | 31.2 (25.9 to 36.5) | 32.9 (27.3 to 38.4) | 31.6 (25.8 to 37.4) | 32.1 (26.9 to 37.4) |
| Limit consumption of sugar sweetened<br>beverages<sup>1,2</sup> | 47.8 (43.7 to 51.9) | 56.0 (50.1 to 61.8) | 39.4 (33.7 to 45.1) | 44.1 (38.7 to 49.4) | 54.2 (47.9 to 60.5) | 45.4 (39.8 to 51.0) | 51.3 (45.3 to 57.2) | 48.7 (42.5 to 54.8) | 46.9 (41.5 to 52.4) |
| Limit the addition of sugar to hot or<br>cold drinks<sup>1</sup> | 34.2 (30.4 to 38.2) | 39.0 (33.3 to 44.7) | 29.2 (23.9 to 34.6) | 31.5 (26.4 to 36.5) | 38.9 (32.8 to 45.0) | 31.3 (26.0 to 36.6) | 38.4 (32.7 to 44.2) | 35.5 (29.5 to 41.5) | 33.0 (27.9 to 38.1) |
| Limit use of instant drink mixes<sup>3</sup> | 13.0 (10.5 to 16.0) | 13.2 (9.3 to 17.2) | 12.8 (9.1 to 16.5) | 11.7 (8.3 to 15.0) | 15.4 (10.8 to 19.9) | 9.5 (6.2 to 12.9) | 18.2 (13.6 to 22.8) | 12.3 (8.2 to 16.4) | 13.6 (9.9 to 17.2) |
| Limit consumption of confectionery<sup>3</sup> | 8.5 (6.5 to 11.1) | 9.7 (6.1 to 13.2) | 7.4 (4.6 to 10.1) | 7.8 (5.0 to 10.6) | 9.8 (6.0 to 13.6) | 6.1 (3.5 to 8.8) | 12.1 (8.2 to 16.1) | 10.1 (6.4 to 13.9) | 7.2 (4.5 to 9.8) |
| Limit consumption of baked goods | 15.7 (12.9 to 18.9) | 16.2 (11.9 to 20.6) | 15.1 (10.9 to 19.2) | 13.7 (9.9 to 17.4) | 19.1 (14.1 to 24.2) | 14.7 (10.7 to 18.6) | 17.2 (12.7 to 21.7) | 18.6 (13.8 to 23.4) | 13.1 (9.4 to 16.9) |
| Buy low sugar alternatives<sup>2,3</sup> | 4.3 (3.0 to 6.1) | 4.8 (2.4 to 7.1) | 3.9 (2.0 to 5.8) | 2.5 (1.1 to 3.9) | 7.5 (4.2 to 10.9) | 1.3 (0.2 to 2.5) | 8.8 (5.4 to 12.2) | 5.1 (2.6 to 7.6) | 3.7 (1.8 to 5.5) |

\* Fijian of Indian descent or other category. <sup>1</sup> Significant difference by sex, <sup>2</sup> Significant difference by age, <sup>3</sup> Significant difference by area, <sup>4</sup> Significant difference by ethnicity.

### Sugar-related attitudes

The majority (84.2% (80.8 to 87.1%)) felt it was very or somewhat important for them to lower sugar in their diet. A higher proportion of women compared to men (89.4% (85.8 to 93%) vs 78.9% (73.8 to 84.0%) and older compared to younger people (88.8% (84.9 to 92.8%) vs 81.6 (77.2 to 85.9%)) reported that it was very or somewhat important to lower sugar intake, **table 2**.

### Sugar-related behaviors

Almost two-thirds of the study population (65% (61.0 to 69.0%)) reported trying to reduce their sugar intake, with just under half (47.8% (43.7 to 51.9%)) trying to limit their consumption of sugar-sweetened beverages. However, 64.6% (60.6 to 68.5%) of participants reported adding sugar to drinks, and 20% (16.8 to 23.5%) were drinking sugar-sweetened beverages at least 5 days a week.

Sugar-related behaviors differed by population subgroups. Significantly more women than men (73.5% (68.2 to 78.8%) vs. 56.6% (50.6 to 62.5%)) and older adults compared to younger adults (74.5% (69.0 to 80.0%) vs. 59.7% (54.3 to 65.1%)) reported trying to reduce their sugar intake. More women than men (56% (50.1 to 61.8%) vs. 39.4% (33.7 to 45.1%)) and older compared to younger people (54.2% (47.9 to 60.5%) vs 44.1% (38.7 to 49.4%)) were limiting their intake of sugar-sweetened beverages, reflected in lower numbers of women and older people who reported drinking sugar-sweetened beverages daily. A higher proportion of older people were also limiting their consumption of processed foods (39.5% (33.3 to 45.6%) vs 27.6% (22.6 to 32.6%)).

## Discussion

From this representative survey of adults in the central division of Fiji, of which a high proportion were living with overweight or obesity and hypertension, we generally found good knowledge of the relationship between excess salt or sugar consumption and key related risk factors such as hypertension and diabetes. Participants reported positive attitudes towards reducing salt and sugar in diets. However, knowledge of recommended salt and sugar intakes was low and reported behaviours did not match positive attitudes towards reduction, particularly regarding processed food consumption, adding salt when cooking, and adding sugar to drinks. Younger participants and men generally had more negative attitudes and reported less healthy behaviours. These findings have implications for designing interventions in Fiji but also other Pacific Island Countries. Multi-strategy interventions, including awareness raising campaigns on how to reduce salt and sugar and supportive changes to the food environment are needed to help Fijians make food choices that align with the desire to eat healthier foods.

While knowledge of diseases linked to excess salt or sugar intake was high, knowledge of salt and sugar recommendations were low, and individual behaviours were not conducive to lowering salt and sugar intake. These findings highlight a need for behaviour change campaigns that raise awareness of the sugar and salt intake recommendations in parallel with supporting people to change specific behaviours. Examples of successful behaviour change campaigns have common features of building on existing knowledge, focusing on context-specific messages and, engaging with communities to understand how to portray certain behaviours, with messages communicated through context specific media channels (to achieve the most reach to the target audiences)^24^. Evidence also suggests messaging that follows a behaviour change framework can be more effective. For example, a systematic review of diet interventions with messaging facilitated by the Social Cognition Model found improvements in consumption of at least one health promoting food group ^25^. Examples from NCD reduction interventions more broadly have found theory informed text messaging, providing health care information directly to participants, have been effective among racially and socioeconomically diverse groups ^26 27^. From our findings, campaigns in Fiji should target younger people and men in particular.

The WHO recommendation for salt intake (a maximum of 5g a day) can be easily depicted, given this is roughly a maximum of 1 teaspoon of salt daily, albeit salt intake can still be hard to identify when it is in the form of salt already in processed foods or meals. Similarly understanding, and benchmarking sugar intake can be difficult for individuals, with WHO recommending a maximum of 10% of total energy intake coming from “free” sugars, yet ideally 5% of energy intake or less^28^. It is hard to know what this looks like in terms of food consumed, and the reference to “free” sugars also requires a level of nutrition knowledge, as the recommendation refers to sugar intake, excluding sugar from fruit, vegetables, and milk^29^.

An alternative approach is to raise awareness of common sources of salt and sugar and promote a “less is best” message. The reported behaviours of adding salt to cooking and sugar to drinks were verified in our previously published findings, where we found that salt in mixed cooked dishes and sugar from hot drinks were key contributors to intake^20^. Therefore, there is a need for behaviour change campaigns to target these behaviours specifically.

Previous studied have demonstrated that the food environment in Fiji and other Pacific Island Countries largely promote unhealthy rather than healthy food choices with respect to salt and sugar^12 30 31^. Impactful behaviour change depends on personal beliefs and a supportive social, environmental, and political context.^32^ Therefore, behaviour change campaigns are most successful when also supported by political and social will to change the environment so that the healthiest (lower in salt and lower in sugar) options are also the most accessible options. This study suggests the food environment in Fiji largely promotes unhealthy rather than healthy food choices with respect to salt and sugar. This is in line with the quantitative evidence of excess consumption of salt and sugar from the 24-hour diet survey^20^ and supports our earlier research which found that processed packaged foods available for sale in Fiji are high in salt and sugar^15 16^. As such targeting interventions at processed packaged foods is warranted to support behaviour change. Multiple “best buy” interventions have been recommended by the World Health Organisation^3^. For example, through salt and sugar targets where maximum levels of salt and sugar in processed packaged foods are set, requiring the food industry to reformulate. Fiji has experience setting voluntary salt targets, however, change to salt levels in the food supply were limited^15^. As such, this experience highlights the need for mandatory targets with stronger governance mechanisms. Fiji also has a sugar-sweetened beverage tax; however, previous research has shown that this tax needs to be higher achieve health impacts ^18 19 33^. Applying a tax to processed packaged foods high in sugar and salt would complement the beverage tax and revenue could be used to subsidise healthier food options such as fruit and vegetables. Our findings suggest such interventions would particularly benefit younger men^20^. Encouragingly, since the completion of the survey, the Fijian government have announced increased taxes on sugary drinks, other processed foods, alcohol, and tobacco in their budget,^34^ and there are supportive actions in the 2024 National NCD strategic plan.

### Strengths and limitations

We used the WHO STEPs survey for salt and followed a similar structure of questions for sugar; this will allow for similar questions to be followed in the future, including in WHO STEPs surveys and in national nutrition surveys. Local trained researchers used culturally sensitive procedures for data collection, including obtaining the endorsement of the government and the community leaders and informed consent from participants, with research assistants who could speak and conduct the survey in languages preferred by participants. We gathered representative data from a rural and an urban community in Fiji. Finally, results have been disseminated to members of the Provincial Council, including the Turaganikoro (village headman), raising awareness of the findings even at the preliminary stage, further increasing the translation and impact of this work. Researchers are currently working with the Ministry of Health to develop and disseminate an animation targeting sugar consumption and the behaviours of adding sugar to hot drinks, with plans to develop further communication materials based on the present findings.

It is also important to consider these findings in light of the study limitations. This was a cross-sectional survey, and as such only captures KAB at a single time point. Two enumeration areas in the Central Division of Fiji were selected, which means findings may not be generalisable to other areas, particularly people living on other islands in Fiji. As with any self-reported information, it is possible that social desirability bias influenced the results^35^, as people may have been more likely to report positive attitudes and behaviours. We also limited our focus to adults (aged 18 years or older) and as such do not have findings for younger populations.

## Conclusion

In conclusion, this study revealed that adults living with multiple NCD risk factors in Fiji have high levels of knowledge regarding the health implications of salt and sugar consumption and are aware of the need to reduce their intake. However, awareness of recommended intake levels was low, and certain behaviours, such as adding salt during cooking and adding sugar to drinks, were barriers to salt and sugar reduction.

To address these challenges, it is imperative for the Fijian government to help bridge the gap between knowledge and practice regarding dietary salt and sugar behaviour. Support is needed to empower individuals to make healthier choices both through individual behaviour change and a supportive food environment. Given the ongoing diet related burden of disease in Fiji, actions to improve salt and sugar knowledge and behaviours are urgently needed.

## Data Availability

All relevant data are within the manuscript and its Supporting information files. The datasets used and/or analysed during the current study are available from the corresponding author on reasonable request.

## Acknowledgments

The authors wish to thank all participants for contributing to the research. The authors would specifically like to thank Fiji’s Ministry of iTaukei Affairs, the village chiefs, and the Ministry Local Governments for their endorsement of working with the communities in Waidamudamu and Deuba, and all the research assistants who worked tirelessly, braving bad weather and ensuring respect for participants, national health, and the research process.

## Author Contributions

### Conceptualization

Jacqui Webster, Colin Bell, Joseph Alvin Santos, Mark Woodward, Kris Rogers, Briar McKenzie, Gade Waqa

## Data curation

Gade Waqa, Anasaini Moala Silatolu, Joseph Alvin Santos, Kris Rogers, Briar McKenzie, Colin Bell, Jacqui Webster

## Formal analysis

Joseph Alvin Santos, Briar McKenzie, Anasaini Moala Silatolu and Gade Waqa

## Investigation

Joseph Alvin Santos, Briar McKenzie, Anasaini Moala Silatolu, Gade Waqa **Methodology:** Colin Bell, Jacqui Webster, Joseph Alvin Santos, Briar McKenzie, Gade Waqa

## Project administration

Briar McKenzie, Anasaini Moala Silatolu, Gade Waqa **Writing – original draft:** Gade Waqa, Briar McKenzie, Colin Bell, Jacqui Webster **Writing – review & editing:** Gade Waqa, Briar McKenzie, Colin Bell, Jacqui Webster, Aliyah Palu, Alvina Deo, Joseph Alvin Santos, Kris Rogers, Erica Reeve

